# Knowledge, Awareness, and Attitudes Toward Epilepsy in Palestine; A Cross-Sectional Study

**DOI:** 10.1101/2025.05.13.25327550

**Authors:** Alhareth M. Amro, Anas K. Assi, Yahya Kayed AbuJwaid, Salahaldeen Deeb, Habeeb H. Awwad, Amro Odeh

## Abstract

**Background:** Epilepsy is a prevalent neurological disorder often surrounded by misinformation and stigma, especially in resource-constrained settings. Despite effective treatments, public misconceptions can hinder diagnosis, treatment adherence, and social inclusion. In Palestine, where cultural beliefs and healthcare access are shaped by complex socio-political factors, understanding public perceptions of epilepsy is vital.

**Objective:** This study aimed to assess the level of knowledge, awareness, and attitudes toward epilepsy among the Palestinian population and to identify sociodemographic factors influencing these domains.

**Methods:** A cross-sectional survey was conducted from January to April 2025 using a structured, self-administered questionnaire distributed online. The sample included 570 Palestinian adults aged 18–65 years. Participants’ knowledge, awareness, and attitudes were evaluated using validated scoring systems. Statistical analyses included chi-square tests and multivariate logistic regression to determine predictors of positive outcomes.

**Results:** Overall, 52.1% of participants demonstrated good knowledge of epilepsy, 49.5% had good awareness, and 82.5% expressed positive attitudes toward individuals with epilepsy. Educational attainment, particularly at the postgraduate level, was the strongest predictor of good knowledge (OR = 5.60), good awareness (OR = 5.60), and positive attitude (OR = 4.38). Male gender was independently associated with lower awareness (OR = 0.59). Despite knowledge gaps, a broadly accepting public stance was observed.

**Conclusion:** While knowledge and awareness of epilepsy remain moderate in the Palestinian population, attitudes are overwhelmingly positive. Educational initiatives targeting knowledge deficits, especially among males and less-educated groups, can leverage this positive social foundation to reduce stigma and improve epilepsy care and inclusion. Tailored public health strategies are essential to promote accurate understanding and effective first-aid responses.

## Introduction

Epilepsy is a chronic neurological condition marked by spontaneous and repeated seizures resulting from irregular electrical discharges in the brain. It can affect individuals across all age groups and arises from multiple factors, including genetic traits, brain trauma, developmental issues, or infections [1,2]. According to the World Health Organization (WHO), epilepsy affects over 50 million people globally, presenting a significant health burden, particularly in resource-constrained regions where access to specialized care and public awareness is often lacking [3].

Despite the availability of effective treatments, epilepsy remains misunderstood in many parts of the world. Cultural misconceptions, including beliefs that seizures are caused by spiritual forces, curses, or contagious illnesses, continue to influence how the condition is perceived. These stigmatizing views often lead to social rejection, discrimination, and a reluctance among individuals to disclose their diagnosis or seek medical help, thereby hindering timely care and treatment adherence [4, 5].

Enhancing public understanding is essential to addressing these issues. Educational outreach helps debunk myths, encourages early diagnosis, and equips community members to offer appropriate support during seizures [6]. Research in countries such as Ethiopia, Turkey, and Lebanon have highlighted major gaps in public knowledge and negative societal attitudes, underscoring the importance of culturally relevant awareness campaigns [7–9].

In the Palestinian context, epilepsy remains poorly understood. Many individuals are unaware of proper first aid procedures during seizures and continue to associate the condition with mental illness or supernatural causes [10]. The ongoing impact of military occupation—through limited healthcare access, disrupted services, and insufficient mental health support—may further contribute to misinformation and stigma [11].

This study aims to evaluate the awareness, knowledge, and attitudes toward epilepsy within the Palestinian population, with the goal of guiding future educational strategies and promoting social inclusion.

## Methodology

### Study Design and Setting

We conducted a cross-sectional survey between January and April 2025 to assess public knowledge, awareness, and attitudes toward epilepsy in the Palestinian population. Data were collected online using a structured, self-administered questionnaire hosted on Google Forms. Snowball sampling was employed via social media platforms (WhatsApp, Facebook, and Instagram) to reach a broad, community-based sample of adult Palestinians aged 18–65 years.

### Participants

Inclusion criteria were: Palestinian residency, age between 18 and 65 years, and ability to complete an online survey in Arabic or English. We excluded healthcare professionals and students enrolled in healthcare disciplines (medicine, pharmacy, nursing, midwifery, laboratory sciences) to avoid knowledge bias. A minimum sample size of 269 was determined a priori using Epi Info™ (expected awareness proportion 77.4%, 95% confidence interval, 5% margin of error); we ultimately enrolled 570 participants.

### Questionnaire Development and Translation

The survey was adapted from validated tools used by Younes et al. (2024) in Lebanon [9]. It comprised five sections: (1) sociodemographic data (age, gender, marital status, education, occupation, income, health insurance, area of residence); (2) medical and social history (chronic conditions, smoking, alcohol use); (3) prior epilepsy education (attendance of lectures/seminars, self-perceived knowledge); (4) knowledge items; and (5) awareness and attitude items. Knowledge items included four binary questions (e.g., “Is epilepsy contagious?”) and four multi-response questions on etiology, clinical manifestations, first-aid measures, and treatment options. Awareness was assessed by five binary items (e.g., having witnessed a seizure, knowing someone with epilepsy). Attitude encompassed 14 statements rated “Yes,” “No,” or “Don’t know” (e.g., willingness to work with persons with epilepsy). The questionnaire was forward-translated into Arabic and back-translated to English by independent bilingual experts, with discrepancies reconciled by a third reviewer to ensure semantic and cultural equivalence.

### Scoring and Reliability

Responses were numerically coded: for binary and attitude items, “Yes” =2, “No” =1, “Don’t know” =0; for multi-response knowledge items, each correct option received 1 point. Total scores ranged 0–16 for knowledge, 0–10 for awareness, and 0–28 for attitude. Cutoffs were set at ≥9 for “good knowledge,” ≥6 for “good awareness,” and ≥17 for “positive attitude,” mirroring the Lebanese study. Internal consistency was evaluated via Cronbach’s α: attitude (α=0.819), knowledge (α=0.634), and awareness (α=0.721).

### Data Management and Statistical Analysis

Data were exported from Google Forms into SPSS v25 (IBM Corp., Armonk, NY). Descriptive statistics (means ± SD, frequencies, and percentages) summarized participant characteristics and scale scores. Bivariate associations between sociodemographic variables and categorical outcomes (good vs. poor knowledge/awareness; positive vs. negative attitude) were examined using chi-square tests; variables with p<0.20 were entered into multivariate models. Three separate logistic regression analyses identified independent predictors of good knowledge, good awareness, and positive attitude. Adjusted odds ratios (ORs) with 95% confidence intervals (CIs) and p-values were reported; statistical significance was set at p<0.05. Model fit was assessed via McFadden’s pseudo-R².

### Ethical Approval

The study protocol, including the survey instrument and informed-consent procedure, was reviewed and approved by the Al-Quds University Research Ethics Committee. Participation was voluntary and anonymous; all respondents provided informed consent electronically before accessing the questionnaire.

## Result

A total of 570 participants from the Palestinian population were included in this study. The majority of respondents were female (54.2%), and the most represented age group was 18–25 years (35.1%), followed by 26–35 years (23.9%). More than half of the participants held a university degree (51.9%), while 21.9% had completed postgraduate studies. Regarding employment status, 38.1% were students and 20.7% were employed in the public sector. Most respondents were non-smokers (81.6%), and nearly 60% had a monthly family income below 6,000₪. Only 41.6% reported having health insurance. The majority resided in central and northern areas of the West Bank (Table 1).

**Table 1.** Sociodemographic Characteristics of Study Participants.

| Variables | n (%) |
| --- | --- |
| Female | 309 (54.2%) |
| Male | 261 (45.8%) |
| 18-25 | 200 (35.1%) |
| 26-35 | 136 (23.9%) |
| 36-45 | 122 (21.4%) |
| 46-55 | 77 (13.5%) |
| 56-65 | 35 (6.1%) |
| Married | 332 (58.2%) |
| Single | 220 (38.6%) |
| Divorced | 10 (1.8%) |
| Widowed | 8 (1.4%) |
| University | 391 (68.6%) |
| Postgraduate | 79 (13.9%) |
| Secondary | 74 (13.0%) |
| Intermediate | 20 (3.5%) |
| Primary | 6 (1.1%) |
| Public servant | 144 (25.3%) |
| Student | 128 (22.5%) |
| Private-sector employees | 102 (17.9%) |
| Housewife | 71 (12.5%) |
| Freelancer | 69 (12.1%) |
| Unemployed | 40 (7.0%) |
| Retired | 16 (2.8%) |
| No | 440 (77.2%) |
| Yes | 130 (22.8%) |
| 3001 - 6000 ₪ | 187 (32.8%) |
| 1500 - 3000 ₪ | 186 (32.6%) |
| Less than 1500 ₪ | 101 (17.7%) |
| More than 6000 ₪ | 96 (16.8%) |
| Governmental | 354 (62.1%) |
| Private | 100 (17.5%) |
| None | 97 (17.0%) |
| UNRWA | 19 (3.3%) |
| Hebron | 236 (41.4%) |
| Salfit | 116 (20.4%) |
| Ramallah and Al-Bireh | 107 (18.8%) |
| Nablus | 65 (11.4%) |
| Jenin | 13 (2.3%) |
| Jerusalem | 10 (1.8%) |
| Palestinian inside the Occupied Palestinian Territory | 7 (1.2%) |
| Bethlehem | 7 (1.2%) |
| Tubas | 4 (0.7%) |
| Tulkarm | 4 (0.7%) |
| Qalqilya | 1 (0.2%) |

Values are expressed as number (percentage).

The analysis of scores revealed that the mean Knowledge Score was 8.5 ± 2.9 (range: 0–16), with 52.1% of participants categorized as having good knowledge. The mean Awareness Score was 5.6 ± 1.8 (range: 0–10), and 49.5% had good awareness. The mean Attitude Score was 20.7 ± 4.9 (range: 0–28), with 82.5% demonstrating a positive attitude toward people with epilepsy. Internal reliability analysis showed a Cronbach’s alpha of 0.819 for the Attitude scale (excellent), 0.634 for the Knowledge scale (acceptable), and 0.393 for the Awareness scale (low).

Chi-square analysis indicated a strong relationship between educational level and all three outcome domains. Educational attainment was significantly associated with knowledge level (p = 0.0006), awareness (p = 0.0016), and attitude (p = 0.0056). No statistically significant associations were observed between knowledge, awareness, or attitude and gender, age, or occupation in bivariate analysis, though some trends were noted in favor of younger participants and those with higher education (Table 2).

**Table 2.** Bivariate Associations Between Sociodemographic Factors and Knowledge, Awareness, and Attitude Levels Toward Epilepsy.

| Characteristic | Knowledge (p) | Awareness (p) | Attitude (p) |
| --- | --- | --- | --- |
| Gender | 0.8218 | 0.1084 | 0.9707 |
| Age | 0.1349 | 0.4703 | 0.8367 |
| Marital Status | 0.7105 | 0.9695 | 0.3902 |
| Educational level | 0.0006* | 0.0016* | 0.0056* |
| Occupation | 0.0795 | 0.1743 | 0.7288 |
| Smoker | 0.4013 | 0.6009 | 0.7439 |
| Family income per month | 0.2001 | 0.2904 | 0.0281* |
| Health insurance | 0.0405* | 0.4680 | 0.2416 |
| Area of residence | 0.2731 | 0.0760 | 0.1642 |

p-values based on chi-square tests. Asterisks (\*) indicate $p < 0.05$ .

Multivariate logistic regression revealed several important independent predictors. In the Awareness model, being male was associated with lower awareness (OR = 0.59; 95% CI: 0.40– 0.88; *p* = 0.0103). In contrast, university education (OR = 2.82; 95% CI: 1.05–7.59; *p* = 0.0402) and postgraduate education (OR = 5.60; 95% CI: 1.79–17.55; *p* = 0.0031) were associated with significantly higher awareness. In the Attitude model, postgraduate education was a strong positive predictor (OR = 4.38; 95% CI: 1.41–13.65; *p* = 0.0107). Full results of these significant associations are summarized in Table 3.

**Table 3.** Multivariate Logistic Regression: Significant Predictors of Good Awareness and Positive Attitudes Toward Epilepsy.

| Model | Predictor | OR | 95% CI | p-value |
| --- | --- | --- | --- | --- |
| Awareness | Gender Male | 0.59 | (0.40 – 0.88) | 0.0103* |
| Awareness | Educational level University | 2.82 | (1.05 – 7.59) | 0.0402* |
| Awareness | Educational level Postgraduate | 5.60 | (1.79 – 17.55) | 0.0031* |
| Attitude | Educational level Postgraduate | 4.38 | (1.41 – 13.65) | 0.0107* |

Odds ratios (ORs) with 95% confidence intervals (CI) are shown. Asterisks (\*) indicate $p < 0.05$ .

A focused logistic regression model using Knowledge Level as the sole dependent variable was also conducted. While none of the predictors reached statistical significance, a trend was observed among participants aged 26–35 years (OR = 1.53; 95% CI: 0.82–2.85; *p* = 0.1818). Male gender was associated with lower odds of good knowledge (OR = 0.79; *p* = 0.2382), but the result was not significant. Full details are presented in Table 4.

**Table 4.** Logistic Regression Analysis of Factors Associated with Good Knowledge About Epilepsy Among Participants.

| Variable | OR (95% CI) | p-value |
| --- | --- | --- |
| Constant | 1.08 (0.33–3.52) | 0.8960 |
| Gender Male | 0.79 (0.53–1.17) | 0.2382 |
| Age 26-35 | 1.53 (0.82–2.85) | 0.1818 |
| Age 36-45 | 1.11 (0.58–2.13) | 0.7449 |
| Age 46-55 | 0.74 (0.36–1.54) | 0.4208 |
| Age 56-65 | 0.96 (0.35–2.60) | 0.9352 |
| Educational level Postgraduate | 2.62 (0.87–7.83) | 0.0857 |
| Educational level Primary | 0.17 (0.02–1.80) | 0.1413 |
| Educational level Secondary | 0.77 (0.28–2.15) | 0.6181 |
| Educational level University | 1.48 (0.57–3.81) | 0.4192 |
| Occupation Housewife | 0.86 (0.40–1.86) | 0.7081 |
| Occupation Private-sector employees | 1.73 (0.88–3.39) | 0.1108 |
| Occupation Public servant | 0.94 (0.49–1.79) | 0.8394 |
| Occupation Retired | 1.44 (0.36–5.74) | 0.6016 |
| Occupation Student | 1.17 (0.56–2.44) | 0.6834 |
| Occupation Unemployed | 0.69 (0.31–1.57) | 0.3810 |

Logistic regression examining predictors of good knowledge level. OR = odds ratio; CI = confidence interval. p-values <0.05 are considered statistically significant.

Finally, a comparative regression table summarizing predictors across all three outcomes Knowledge, Awareness, and Attitude was developed. This table highlights that education level, especially at the postgraduate level, is a consistently strong and significant predictor across all models. Meanwhile, male gender remains a risk factor for lower awareness (Table 5).

**Table 5.**
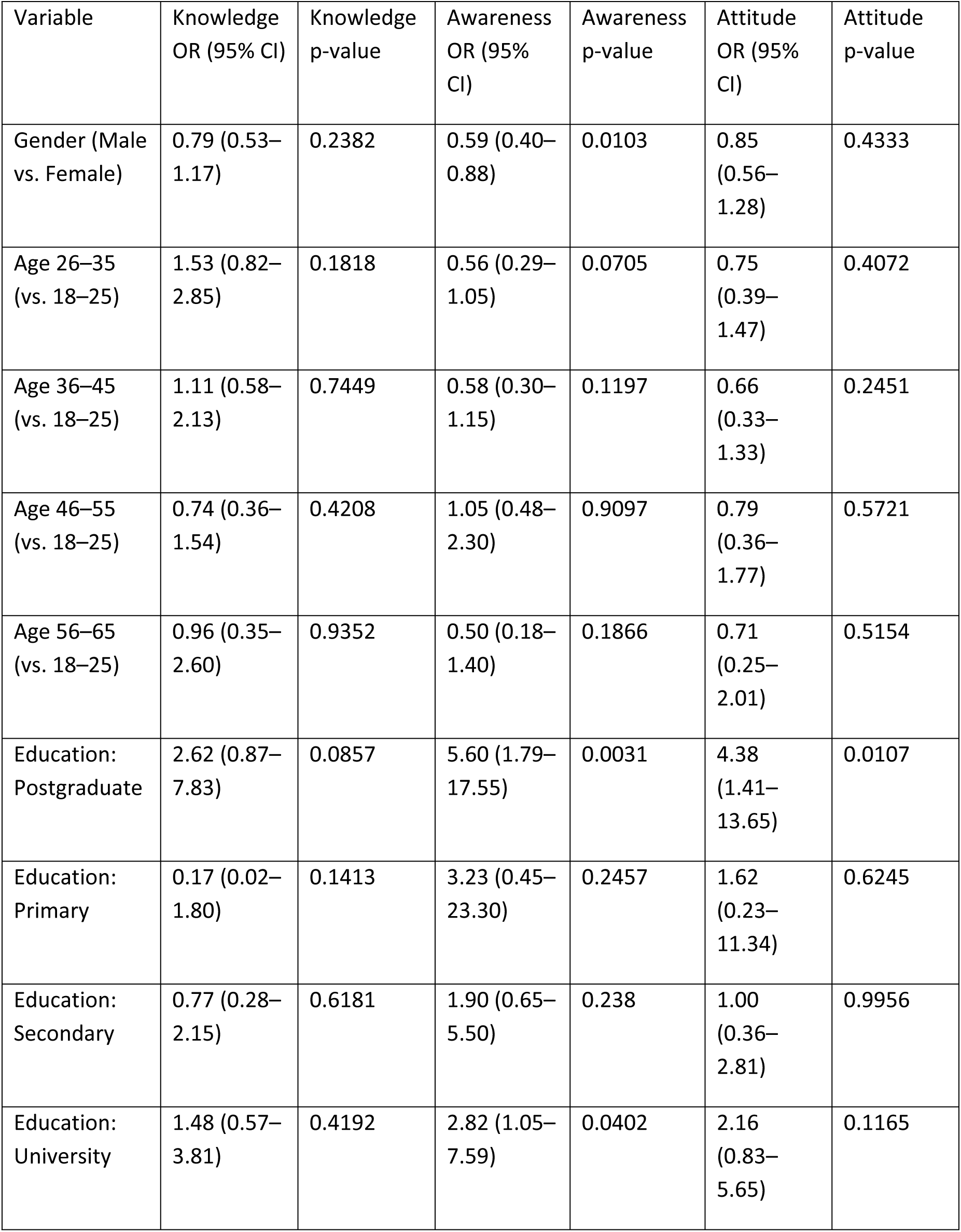
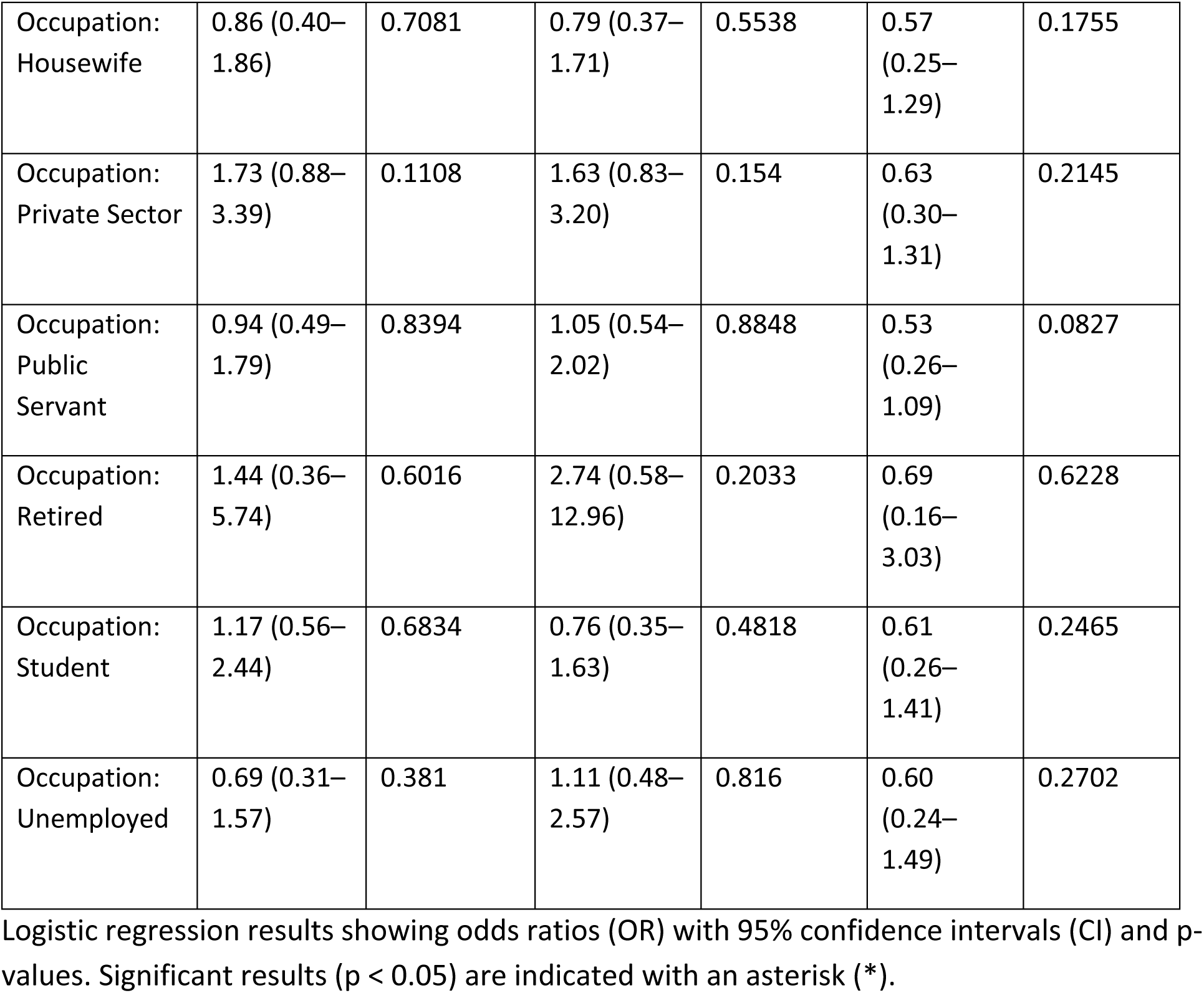
Factors Associated with Knowledge, Awareness, and Attitude Toward Epilepsy Among Participants.

## Discussion

This study provides a comprehensive overview of public knowledge, awareness, and attitudes toward epilepsy in the Palestinian population. The findings reveal a nuanced picture: while just over half of participants demonstrated good knowledge (52.1%; mean score 8.5 ± 2.9) and awareness (49.5%; mean score 5.6 ± 1.8) of epilepsy, an overwhelming majority (82.5%; mean score 20.7 ± 4.9) exhibited a positive attitude toward individuals living with the condition.

Educational attainment, particularly at the postgraduate level, emerged as the most consistent and significant predictor across all three outcome domains. Notably, male gender was associated with lower awareness levels (OR = 0.59; 95% CI: 0.40–0.88; p = 0.0103), highlighting a gender disparity that warrants attention.

These findings align partially with regional and international studies. A recent Lebanese survey reported significantly higher rates of good knowledge (87.4%) and awareness (70.1%), though the proportion of participants with positive attitudes (88%) closely matched our findings [9]. The discrepancy in knowledge and awareness may reflect differences in public health infrastructure, the reach of continuing education programs, or cultural factors influencing information dissemination. Despite only moderate knowledge levels in our sample, most participants correctly identified that epilepsy is not contagious and recognized key clinical features.

However, less than one-quarter knew it is incurable, underscoring specific misconceptions that should be addressed in future educational initiatives.

Other countries in the Middle East show varied patterns. A Jordanian study found that only 35.3% of participants had good knowledge of epilepsy, although 63.3% reported favorable attitudes [12]. In Saudi Arabia, persistent cultural misconceptions were evident; over half of the respondent’s attributed epilepsy to supernatural causes such as jinn, and only 30% correctly identified first-aid measures during a seizure [13]. Compared to these countries, the Palestinian population demonstrated relatively better medical knowledge and more supportive social attitudes, despite enduring cultural myths.

In Bahrain, public awareness was very high (95.6%), but stigma remained a concern. For example, 27.5% of participants opposed marriage with individuals who have epilepsy [14]. In contrast, Palestinian participants, while exhibiting lower awareness levels, showed greater acceptance and positive attitudes. This contrast underscores the importance of evaluating both cognitive understanding and social perceptions in assessing epilepsy literacy.

Outside the MENA region, similar trends are observed. A 2007 study in Turkey reported moderate public awareness, with 45.2% showing good knowledge and 55.5% demonstrating positive attitudes [15]. In Cameroon’s Akwaya District, a 2009 study revealed high levels of misinformation; 25.5% believed epilepsy was contagious, and 33.3% viewed it as a form of mental illness. Moreover, 38.1% felt people with epilepsy should not marry, and 28.6% believed they should not have children. Education again emerged as a strong predictor of better knowledge and attitudes [16]. Compared to these contexts, the Palestinian population reflected better outcomes in both knowledge and attitude, albeit with continued misconceptions among certain subgroups.

A 2024 study of Chinese university students found nearly universal awareness (94.7%), especially among medical students, and high levels of accurate first-aid knowledge [17]. In contrast, a meta-analysis of Ethiopian studies (2023) reported that only about 47% of respondents demonstrated good knowledge and a similarly low rate of favorable attitudes [7]. These disparities illustrate how resource availability, education infrastructure, and cultural narratives influence epilepsy perceptions across different regions.

Educational attainment was the single strongest determinant across all domains in our study. Participants with postgraduate education were more than five times as likely to have good awareness (OR = 5.60; 95% CI: 1.79–17.55; p = 0.0031) and nearly five times as likely to hold positive attitudes (OR = 4.38; 95% CI: 1.41–13.65; p = 0.0107) compared to those without formal higher education. University graduates also had over double the odds of good awareness (OR = 2.82; 95% CI: 1.05–7.59; p = 0.0402). This trend is mirrored in several international studies. In Lebanon, widespread health education initiatives were associated with higher knowledge and awareness levels [9]. Similarly, the Chinese study highlighted the role of academic exposure, particularly in health-related fields, in shaping informed perspectives [17]. Conversely, countries with limited educational outreach, such as Jordan [12], demonstrated lower levels of epilepsy knowledge.

Cultural beliefs also play a crucial role. In Saudi Arabia, traditional explanations such as jinn possession persist despite medical advances [13]. Bahrain, although more medically informed, still reported significant stigma [14]. These findings emphasize that improving knowledge does not automatically eliminate negative attitudes. Interestingly, our study found the reverse trend—a relatively informed but highly empathetic public—suggesting that social norms in Palestine may be shifting toward greater inclusivity. Furthermore, nearly half of respondents still believed that people with epilepsy should not drive—a view that, while sometimes clinically justified, may also reflect lingering stigma. Nevertheless, 75% of participants supported full social integration in employment and marriage, aligning with broader regional improvements in public attitudes.

Healthcare system limitations also influence public understanding. In countries like Lebanon and China, better-organized health sectors and integrated public health campaigns have fostered greater epilepsy literacy. In contrast, the Palestinian context, characterized by political instability, restricted healthcare access, and limited funding for long-term awareness programs, likely contributes to the observed gaps in knowledge and awareness. Nonetheless, the high rate of positive attitudes indicates a societal readiness to support individuals with epilepsy if provided with accurate information and resources.

This study has several strengths. It is among the first large-scale surveys to assess epilepsy-related knowledge, awareness, and attitudes in Palestine. The use of a validated questionnaire allows for meaningful comparison with international data. The sample size was robust, and internal reliability metrics were within acceptable ranges for most scales—acceptable for knowledge (α = 0.634) and excellent for attitude (α = 0.819), though low for awareness (α = 0.393), likely due to the limited number of items. This mirrors the Lebanese experience and highlights the need to refine awareness measures in future research. The cross-sectional design limits causal inference, and self-reported data may introduce recall or social desirability bias. Additionally, online sampling likely skewed the participant pool toward younger and more educated individuals, potentially overestimating knowledge levels. The absence of a qualitative component further limits insight into the deeper cultural and emotional underpinnings of public attitudes.

Future research should address these gaps by employing community-based sampling to include underrepresented groups such as older adults and rural populations. Mixed-methods approaches incorporating qualitative interviews or focus groups could uncover the nuanced beliefs and experiences that shape public perceptions of epilepsy. Additionally, longitudinal studies are recommended to evaluate the effectiveness of public education campaigns and to monitor changes in attitudes and knowledge over time.

The observed gap between moderate knowledge and strong positive attitudes presents a unique public health opportunity. Positive social attitudes provide a fertile ground for targeted educational interventions aimed at correcting misconceptions and improving practical knowledge, particularly regarding first-aid measures and treatment adherence. Public health efforts should prioritize mass media campaigns, school-based health education, and community engagement through religious and local leaders. Supporting families of individuals with epilepsy through counseling and peer networks can further reduce stigma and promote inclusion.

## Conclusion

This study highlights a complex but hopeful picture of epilepsy perceptions in Palestine. While knowledge and awareness remain moderate and marked by specific misconceptions, public attitudes toward individuals with epilepsy are largely positive and inclusive. Education, particularly at higher levels, stands out as the strongest predictor of improved understanding and supportive attitudes. These findings underscore the critical need for well-designed, culturally sensitive educational interventions that target gaps in knowledge and address enduring myths.

Leveraging existing positive attitudes through targeted health education and media outreach can pave the way for greater social inclusion and better health outcomes for people with epilepsy. By building on this supportive societal foundation, future public health initiatives can drive meaningful change in how epilepsy is understood and managed in Palestinian communities.

## Data Availability

All relevant data are within the manuscript and its Supporting Information files.

## Acknowledgements

None.

## Data availability

The datasets used and/or analyzed during the current study are available from the corresponding author on reasonable request.

## Ethics approval and consent to participate

All procedures performed in this study involving human participants complied with the institutional and/or national research committee ethical standards and the 1964 Helsinki declaration and subsequent amendments or equivalent ethical standards. The study was designed and conducted in accordance with the ethical principles established by Al-Quds University.

Therefore, ethical approval was obtained from the Institutional Review Board Committee, Al-Quds University. Written informed consent was obtained from all the participants for the participation of this study and accompanying images. A copy of the written consent is available for review by the Editor-in-Chief of this journal on request.

## Clinical trial number

Not applicable.

## Consent for publication

The manuscript contains no images or videos—not applicable.

## Contributions

All authors: Alhareth M. Amro, Anas K. Assi, Yahya Kayed AbuJwaid, Salahaldeen Deeb, Habeeb H. Awwad, and Amro Odeh contributed equally to this work. Each author was involved in the conception, design, analysis, and interpretation of the research, as well as drafting and revising the manuscript. All authors have participated in writing the manuscript and reviewed the literature. All authors contributed to revision of the manuscript. All authors read and approved the final manuscript.

## References

1. Milligan TA. Epilepsy: a Clinical Overview. The American Journal of Medicine. 2021;134(7):840–847. doi:10.1016/j.amjmed.2021.01.038

2. Anwar H, Khan QU, Nadeem N, Pervaiz I, Ali M, Cheema FF. Epileptic seizures. Discoveries. 2020;8(2):e110. doi:10.15190/d.2020.7 World Health Organization: WHO. Epilepsy. Published February 7, 2024. https://www.who.int/news-room/fact-sheets/detail/epilepsy

3. Yeni K. Stigma and psychosocial problems in patients with epilepsy. Exploration of Neuroscience. 2023;2(6):251–263. doi:10.37349/en.2023.00026

4. Sen A, Newton CR, Ngwende G. Epilepsy in low- to middle-income countries. Current Opinion in Neurology. Published online February 10, 2025. doi:10.1097/wco.0000000000001350

5. Min A, Miller WRT, Connelly K, Nippert-Eng C, Lin HC, Shih PC. Enhancing epilepsy awareness and cooperative care in elementary and middle schools. Proceedings of the ACM on Human-Computer Interaction. 2024;8(CSCW1):1–25. doi:10.1145/3641003

6. Woldegeorgis BZ, Anjajo EA, Korga TI, et al. Ethiopians’ knowledge of and attitudes toward epilepsy: A systematic review and meta-analysis. Frontiers in Neurology. 2023;14. doi:10.3389/fneur.2023.1086622

7. Kayar V, Sahin MK. Adults’ knowledge levels of and attitudes toward epilepsy: a cross-sectional study in Samsun Türkiye. Deleted Journal. 2024;29(1):87–96. doi:10.54029/2024mzr

8. Younes S, Chahine B, Hanna V, et al. Public awareness, knowledge, and attitude toward epilepsy in Lebanon: a cross-sectional study. Frontiers in Neurology. 2024;15. doi:10.3389/fneur.2024.1480960

9. Shawahna R. Epilepsy knowledge and attitudes: A large observational study among the Palestinian general public. Heliyon. 2023;10(1):e23707. doi:10.1016/j.heliyon.2023.e23707

10. Jabali O, Ayyoub AA, Jabali S. Navigating health challenges: the interplay between occupation-imposed movement restrictions, healthcare access, and community resilience. BMC Public Health. 2024;24(1). doi:10.1186/s12889-024-18817-y

11. Abuhamdah SMA, Naser AY, Abualshaar MAR. Knowledge of and Attitude towards Epilepsy among the Jordanian Community. Healthcare. 2022;10(8):1567. doi:10.3390/healthcare10081567

12. AlHarbi FA, Alomari MS, Ghaddaf AA, Abdulhamid AS, Alsharef JF, Makkawi S. Public awareness and attitudes toward epilepsy in Saudi Arabia: A systematic review and meta-analysis. Epilepsy & Behavior. 2021;124:108314. doi:10.1016/j.yebeh.2021.108314

13. Elmazny A, Alzayani S, Shehata MH, Magdy R. Knowledge, awareness, and attitudes towards epilepsy among elementary schoolteachers in the Kingdom of Bahrain. European Journal of Paediatric Neurology. 2023;47:13–17. doi:10.1016/j.ejpn.2023.08.001

14. Demirci S, Dönmez CM, Gündoğar D, Baydar ÇL. Public awareness of, attitudes toward, and understanding of epilepsy in Isparta, Turkey. Epilepsy & Behavior. 2007;11(3):427–433. doi:10.1016/j.yebeh.2007.08.005

15. Njamnshi AK, Tabah EN, Yepnjio FN, et al. General public awareness, perceptions, and attitudes with respect to epilepsy in the Akwaya Health District, South-West Region, Cameroon. Epilepsy & Behavior. 2009;15(2):179–185. doi:10.1016/j.yebeh.2009.03.013

16. Zhao T, Zhang X, Cui X, et al. Awareness, attitudes and first aid knowledge of epilepsy among university students – A cross-sectional study in Henan Province, China. Epilepsy Research. 2024;201:107315. doi:10.1016/j.eplepsyres.2024.107315

